# UV-induced Mutations are Associated with Melanoma-Specific Mortality in Stage II/III Melanomas from the InterMEL study

**DOI:** 10.64898/2026.11.18.26368442

**Authors:** Adam Z. Reynolds, Li Luo, Ronglai Shen, Arshi Arora, Irene Orlow, Klaus J. Busam, Cecilia Lezcano, Tim K. Lee, Eva Hernando, Ivan Gorlov, Christopher Amos, Marc S. Ernstoff, Venkat Seshan, Anne E. Cust, James S. Wilmott, Graham Mann, Ismael A. Vergara, Eduardo Nagore, Pauline Funchain, Jennifer Ko, Sharon N. Edmiston, Kathleen Conway, Paul B. Googe, David Ollila, Jeffrey E. Lee, Judy R. Rees, Cheryl L. Thompson, Meg Gerstenblith, Marcus Bosenberg, Bonnie E. Gould Rothberg, Iman Osman, Yvonne Saenger, Matthew Schwartz, Tawny Boyce, Sheri L. Holmen, Shaofeng Yan, Elise Brunsgaard, Paul Bogner, Pei Fen Kuan, Colin B. Begg, Nancy E. Thomas, Marianne Berwick, InterMEL

## Abstract

The prognostic significance of tumor mutational burden (TMB) in melanoma remains controversial. One potential reason for the mixed results is that total TMB treats all mutations as biologically equivalent, despite evidence that different types of mutations contribute differently to a tumor’s neo-antigenic profile. In particular, UV-induced and non-UV mutations may have distinct effects on tumor behavior and outcomes, but this hypothesis has been studied primarily in advanced-stage disease. We analyzed 656 stage II/III tumors from the InterMEL study to evaluate whether UV TMB and non-UV TMB are associated with melanoma-specific mortality and clinicopathologic factors. Higher UV TMB was associated with improved outcomes, while non-UV TMB was not associated with mortality. These findings suggest that an association between total TMB and outcome may be driven disproportionately by mutations associated with UV mutagenesis. Although this has previously been reported in advanced melanomas, our work demonstrates that the prognostic relevance of UV TMB extends to stage II/III tumors typically found in the clinical context. Whether this relationship reflects differences in tumor neo-antigenicity, immune recognition, or other biological properties remains to be determined.

**SIGNIFICANCE:** It has long been theorized that mutation counts in melanoma tumors could be prognostic for immune responses and survival outcomes. However, mixed results have made it difficult to implement tumor mutational burden (TMB) as a biomarker. Here we distinguish UV-induced and non-UV mutations, which are expected to have different effects on disease processes. Our analysis shows that UV-induced mutations have a stronger association with better outcomes, suggesting that UV induced mutation counts may be a better and more reliable prognostic biomarker than total mutation counts. The potential clinical value of a UV TMB-based biomarker should be explored further.

## INTRODUCTION

The ability to distinguish aggressive melanomas from more indolent tumors is a critical research priority with importance for clinical practice. There is a need for accurate prognostic biomarkers to identify patients with high-risk disease in earlier stages (II and III), as 10-year overall survival as low as 75% for stage IIC disease^1^. Along with recent trials of adjuvant anti-PD-1 immunotherapies showing a survival benefit for patients with stage II disease^2,3^, there is the urgent need to balance the potential benefits of these treatments against the risk of adverse events^4,5^. Tumor mutational burden (TMB), the number of somatic alterations per megabase, is an important characteristic of melanoma (as well as other cancers). However, to date, its role in the prognosis of melanoma is poorly understood. TMB is commonly believed to proxy a tumor’s neo-antigenic profile, which influences the immune system’s ability to identify and eradicate cancer cells. Thus, tumors with high TMB are thought to be more recognizable to the immune system, which may improve natural immune responses and increase the chances of a positive response to immune checkpoint therapies^6^.

There is an extensive literature debating the effects of TMB on survival and treatment responses in melanoma, with some finding high TMB to predict better survival^7–9^, some finding no effect^10–12^, and at least one finding worse survival^13^. Additionally, a number of papers find that the effect of TMB on survival depends on its interaction with particular regimes of therapy^11,14,15^ or that it predicts local, but not metastatic, recurrence^16^. These inconsistent associations between total TMB and outcomes may reflect competing mutational profiles with biologically distinct consequences for the neo-antigenic signatures of tumor cells or other disease processes^17–19^. Several investigators have suggested that it may be specifically the UV-induced component of TMB (UV TMB) that stimulates immunogenic factors, leading to better survival^20–24^. However, prior studies have been limited to advanced-stage tumors^20,22^. It remains unclear whether UV TMB is associated with prognosis in the natural history of early-stage tumors or whether there is potential value for UV TMB to contribute to decision-making in the clinical context.

Here we use a large sample of stage II/III primary melanomas from the InterMEL project to investigate the separate contributions of UV TMB and non-UV TMB to tumor pathology and prognosis. We hypothesize that UV TMB and non-UV TMB reflect different mutational processes and thus will differently associate with disease outcomes.

## METHODS

InterMEL is an international collaborative study designed to explore the genomic landscape of stage II/III primary cutaneous melanoma^25^. The InterMEL study consists of 15 participating institutions from the United States, Australia and Spain. Using a case-control design, we analyzed 656 AJCC 8^th^ edition Stage II/III primary melanomas diagnosed between January 1998 and December 2015 from patients untreated with adjuvant therapies who had either died from melanoma within five years of diagnosis (cases) or were alive and free of disease for at least 5 years (controls). Clinicopathological characteristics were reviewed by expert dermatopathologists (KB, CL) at Memorial Sloan-Kettering Cancer Center. Tumor DNA were sequenced for mutational profiles using the MSK-IMPACT™ panel that involves sequencing of 468 major cancer genes^25,26^ and filtering common germline polymorphisms found in normal DNA. For 217 samples with tumor only sequencing, we applied the Glitter (<u>G</u>ermline variant filter) method that distinguishes somatic variants from germline variants in the absence of normal samples^27^. Full laboratory protocols are described in Orlow et al.^28^. Each institution obtained tumors from individuals with informed consent and IRB approval; the entire study was approved by the University of New Mexico Institutional Review Board (HRRC 16-389). All data are de-identified and STROBE guidelines have been followed. All data used in this analysis are available on dbGaP, at accession number phs003099.v1.p1.

Summary statistics including median, quartiles, counts and percentages were used to describe the characteristics of the demographic, clinical and pathological factors, and the genomic mutations identified. Wilcoxon rank-sum tests were used to compare continuous variables. Pearson’s Chi-squared and Fisher’s exact tests were performed to compare categorical variables between the case and control outcome groups. TMB was defined as the number of non-synonymous mutations per megabase sequenced. The UV-induced component of TMB (UV TMB) was estimated by counting mutations that were C>T and CC>TT transitions. Non-UV mutations were identified as those that did not meet these criteria. Medians and inter-quartile ranges were taken from raw TMB values, while all models used log-transformed measures of TMB. Associations between clinicopathologic variables and measures of TMB were tested using linear regression models, adjusting for age and sex. To test the association of TMB measures with mortality outcomes, we used multi-variable logistic regression to examine the effects of UV TMB and non-UV TMB, and estimated odds ratios and 95% confidence intervals with global p-values.

## RESULTS

Patients (n=656) used in the analysis had a median age at diagnosis of 64, were more likely to be male (64%) and diagnosed in stage II (54%) than stage III (46%). The tumors had a median Breslow depth of 4.00mm (0.98mm – 47.00mm), were more likely to be ulcerated (61%), score positively for mitoses (95%), have a non-brisk TIL grade (87%) [brisk (4%), absent (9%)], and a range of solar elastosis [present (46%), mild/moderate (34%), severe (20%)]. Roughly 56% of patients lived longer than 5 years, while 44% died in less than 5 years.

### Clinicopathologic Features

Table 1 summarizes UV TMB, non-UV TMB, and total TMB by clinicopathologic features. Most tumors had greater UV TMB, with UV mutations making up a median of 74% (IQR: 58% – 83%) of all mutations. Males had higher counts of UV mutations than females (*β* = 0.36, *p* < 0.001) and patients in both the 60-79 and 80+ age groups had more mutations of both kinds compared to patients in the under 40 group (60-79: *β* = 0.48, *p* = 0.001; 80+: *β* = 0.53, *p* = 0.002). Tumors on the Head, Neck, and Arms had greater UV TMB (H&N: *β* = 0.52, *p* < 0.001; Arms: *β* = 0.45, *p* < 0.001) and total TMB (H&N: *β* = 0.44, *p* < 0.001; Arms: *β* = 0.37, *p* = 0.001) than tumors on the trunk, while tumors on the legs had lower counts of both UV (*β* = −0.58, *p* < 0.001) and non-UV (*β* = −0.39, *p* < 0.001) mutations as well as Total TMB (*β* = −0.56, *p* < 0.001). Breslow thickness, ulceration, and TIL grade were not associated with overall TMB, UV TMB, or non-UV TMB. Mitoses were associated with non-UV TMB (*β* = 0.43, *p* = 0.004), but not with overall TMB or UV TMB. Perhaps surprisingly, the presence of either mild/moderate or severe solar elastosis were strongly associated with both UV (M/M: *β* = 0.69, *p* < 0.001; Severe: *β* = 1.14, *p* < 0.001) and non-UV mutations (M/M: *β* = 0.41, *p* < 0.001; Severe: *β* = 0.51, *p* < 0.001), as well as with total TMB (M/M: *β* = 0.66, *p* < 0.001; Severe: *β* = 1.02, *p* < 0.001).

**Table 1.** Associations of UV TMB, non-UV TMB, and Total TMB with tumor clinicopathologic characteristics.

|  | UV TMB |  |  |  | Non-UV TMB |  |  |  | Total TMB |  |  |  |
|---|---|---|---|---|---|---|---|---|---|---|---|---|
| Med % (IQR) of Mutations | 74 (58 - 83) |  |  |  | 26 (17 - 42) |  |  |  | 100 |  |  |  |
|  | Median TMB | IQR | Beta<sup>a</sup> | p-value | Median TMB | IQR | Beta<sup>a</sup> | p-value | Median TMB | IQR | Beta<sup>a</sup> | p-value |
| Age at Dx |  |  |  |  |  |  |  |  |  |  |  |  |
| < 40 | 7.27 | (3.64 - 12.73) | Ref. | — | 2.73 | (1.82 - 5.45) | Ref. | — | 11.36 | (4.77 - 16.36) | Ref. | — |
| 40 - 59 | 10 | (4.55 - 18.86) | 0.25 | 0.153 | 3.64 | (1.82 - 8.18) | 0.15 | 0.202 | 14.55 | (6.36 - 25.45) | 0.21 | 0.156 |
| 60 - 79 | 12.73 | (5.45 - 30.91) | 0.55 | 0.001 | 5.45 | (2.73 - 9.09) | 0.26 | 0.019 | 17.27 | (9.09 - 39.09) | 0.48 | 0.001 |
| 80+ | 11.82 | (5.68 - 31.82) | 0.59 | 0.003 | 5.91 | (2.05 - 10.00) | 0.33 | 0.012 | 20.91 | (8.41 - 41.36) | 0.53 | 0.002 |
| Sex |  |  |  |  |  |  |  |  |  |  |  |  |
| Female | 8.18 | (3.64 - 16.36) | Ref. | — | 3.64 | (1.82 - 8.18) | Ref. | — | 12.73 | (5.45 - 23.64) | Ref. | — |
| Male | 11.82 | (5.45 - 27.27) | 0.36 | <0.001 | 4.55 | (2.73 - 9.09) | 0.12 | 0.085 | 17.27 | (9.09 - 35.45) | 0.28 | 0.001 |
| Site |  |  |  |  |  |  |  |  |  |  |  |  |
| Trunk | 10 | (5.45 - 17.73) | Ref. | — | 4.55 | (2.73 - 7.27) | Ref. | — | 15.45 | (9.09 - 23.64) | Ref. | — |
| Head & Neck | 21.82 | (7.27 - 42.73) | 0.52 | <0.001 | 6.36 | (3.64 - 10.91) | 0.18 | 0.04 | 27.27 | (11.82 - 52.73) | 0.44 | <0.001 |
| Arms | 15.91 | (8.18 - 34.55) | 0.45 | <0.001 | 5 | (2.73 - 10.91) | 0.15 | 0.114 | 21.82 | (11.82 - 45.45) | 0.37 | 0.001 |
| Legs | 5.45 | (1.82 -12.73) | -0.58 | <0.001 | 2.73 | (0.91 - 5.45) | -0.39 | <0.001 | 9.09 | (3.64 - 19.77) | -0.56 | <0.001 |
| AJCC8 Stage |  |  |  |  |  |  |  |  |  |  |  |  |
| IIA | 10 | (4.55 - 22.73) | Ref. | — | 3.64 | (1.82 - 8.18) | Ref. | — | 15.45 | (6.82 - 29.09) | Ref. | — |
| IIB | 14.55 | (7.27 - 33.64) | 0.28 | 0.033 | 5.45 | (2.73 - 10.00) | 0.2 | 0.043 | 19.09 | (10.91 - 43.18) | 0.28 | 0.026 |
| IIC | 17.27 | (7.27 - 39.09) | 0.31 | 0.056 | 7.27 | (3.64 - 11.82) | 0.35 | 0.003 | 25 | (11.14 - 48.86) | 0.36 | 0.017 |
| IIIA | 10 | (6.36 - 14.55) | -0.13 | 0.582 | 2.73 | (1.82 - 5.45) | -0.12 | 0.508 | 11.82 | (8.18 - 21.82) | -0.12 | 0.603 |
| IIIB | 8.64 | (4.55 - 18.18) | -0.07 | 0.704 | 3.64 | (1.14 - 8.64) | 0.09 | 0.486 | 12.27 | (6.36 - 31.14) | -0.03 | 0.852 |
| IIIC | 9.09 | (3.64 - 17.27) | -0.19 | 0.150 | 3.64 | (1.82 – 6.36) | -0.04 | 0.642 | 12.73 | (7.27 – 25.45) | -0.13 | 0.278 |
| IIID | 6.36 | (1.82 – 10.00) | -0.57 | 0.011 | 3.64 | (1.82 – 6.36) | -0.11 | 0.512 | 10 | (4.55 – 15.45) | -0.45 | 0.033 |
| Breslow Thickness (mm) |  |  |  |  |  |  |  |  |  |  |  |  |
| < 1.0 | 7.27 | (7.27 – 7.27) | Ref. | — | 2.73 | (2.73 – 2.73) | Ref. | — | 10 | (10.00 – 10.00) | Ref. | — |
| 1.1 – 2.0 | 10 | (5.00 – 16.36) | -0.34 | 0.765 | 3.64 | (1.82 – 7.27) | 0 | 0.995 | 14.55 | (7.27 – 22.27) | -0.19 | 0.855 |
| 2.1 – 4.0 | 11.82 | (4.55 – 25.68) | -0.24 | 0.831 | 4.55 | (1.82 – 9.09) | 0.14 | 0.868 | 16.36 | (7.95 – 35.45) | -0.06 | 0.951 |
| 4.1+ | 10 | (4.55 – 24.55) | -0.34 | 0.76 | 4.55 | (1.82 – 9.09) | 0.06 | 0.944 | 14.55 | (8.18 – 31.82) | -0.16 | 0.879 |
| Ulceration |  |  |  |  |  |  |  |  |  |  |  |  |
| Present | 10 | (4.55 – 22.50) | Ref. | — | 3.64 | (1.82 – 8.18) | Ref. | — | 14.55 | (8.18 – 28.86) | Ref. | — |
| Absent | 10.91 | (4.55 – 26.36) | -0.01 | 0.923 | 4.55 | (1.82 – 9.09) | 0.02 | 0.738 | 16.36 | (8.18 – 34.55) | 0 | 0.965 |
| Nodes |  |  |  |  |  |  |  |  |  |  |  |  |
| N0 | 12.73 | (6.14 – 32.05) | Ref. | — | 5.45 | (2.73 – 10.00) | Ref. | — | 18.64 | (9.09 – 41.14) | Ref. | — |
| N1 | 10 | (4.55 – 18.18) | -0.31 | 0.004 | 3.64 | (1.82- 7.05) | -0.2 | 0.011 | 14.09 | (7.27 – 26.36) | -0.29 | 0.003 |
| N2 | 9.09 | (3.64 – 16.36) | -0.42 | 0.005 | 3.64 | (1.82 – 7.27) | -0.17 | 0.12 | 12.73 | (5.45 – 21.82) | -0.33 | 0.019 |
| N3 | 7.27 | (1.82 – 12.73) | -0.61 | <0.001 | 3.64 | (2.05 – 6.36) | -0.2 | 0.088 | 10.91 | (4.77 – 20.45) | -0.51 | <0.001 |
| Mitoses |  |  |  |  |  |  |  |  |  |  |  |  |
| Absent | 7.27 | (3.86 – 12.05) | Ref. | — | 2.27 | (1.59 – 3.64) | Ref. | — | 10 | (5.23 – 14.77) | Ref. | — |
| Present | 10.91 | (4.55 – 24.32) | 0.14 | 0.492 | 4.55 | (1.82 – 9.09) | 0.43 | 0.004 | 16.36 | (8.18 – 32.73) | 0.26 | 0.177 |
| Solar Elastosis |  |  |  |  |  |  |  |  |  |  |  |  |
| None | 7.27 | (2.73 – 12.73) | Ref. | — | 2.73 | (1.82 – 6.36) | Ref. | — | 10.91 | (4.55 – 19.09) | Ref. | — |
| Mild/Moderate | 12.73 | (7.27 – 27.27) | 0.69 | <0.001 | 5.45 | (2.73 – 10.00) | 0.41 | <0.001 | 19.09 | (10.91 – 37.27) | 0.66 | <0.001 |
| Severe | 25.45 | (9.09 – 45.45) | 1.14 | <0.001 | 7.27 | (3.41 – 11.82) | 0.51 | <0.001 | 32.73 | (14.55 – 56.59) | 1.02 | <0.001 |
| Cannot Assess | 10 | (5.91 – 25.45) | 0.52 | <0.001 | 4.55 | (2.73 – 9.09) | 0.29 | 0.006 | 15.45 | (9.09 – 35.00) | 0.51 | <0.001 |
| TIL grade |  |  |  |  |  |  |  |  |  |  |  |  |
| Absent | 11.36 | (4.31 – 22.73) | Ref. | — | 4.55 | (1.82 – 9.09) | Ref. | — | 15.91 | (6.82 – 29.77) | Ref. | — |
| Non-Brisk | 10.9 | (4.55 – 24.55) | 0.02 | 0.916 | 4.55 | (1.82 – 9.09) | 0 | 0.976 | 15.45 | (8.18 – 32.73) | 0.04 | 0.796 |
| Brisk | 10 | (5.91 – 21.36) | 0.09 | 0.747 | 5.45 | (2.27 – 8.18) | 0.02 | 0.916 | 16.36 | (9.55 – 28.64) | 0.12 | 0.631 |

Note: UV TMB estimated as the count of C>T and CC>TT transitions per sequenced megabase; non-UV TMB estimated as the count of other mutations per sequenced megabase.
<sup>a</sup> Beta estimates are from ordinary linear regressions predicting log-transformed UV TMB, non-UV TMB, and total TMB. All models control for age and sex.
Abbreviations: Med. = Median TMB, IQR = Inter-Quartile Range, Ref. = Reference Category, TIL = Tumor Infiltrating Lymphocytes

### Melanoma-Specific Mortality

A multivariable logistic regression model controlling for other clinicopathologic factors showed that UV TMB was significantly associated with melanoma mortality (OR = 0.70, 95% CI: 0.53 – 0.90)(Table 2). Specifically, the presence of more UV-induced mutations reduces the odds of mortality by 30% per log unit of TMB. In contrast, non-UV TMB showed no association with melanoma-specific mortality (OR = 0.97, 95% CI: 0.70 – 1.35).

**Table 2.** Multivariable logistic regression models testing the association of total TMB, UV TMB, and non-UV TMB with 5-year mortality, controlling for clinicopathologic factors.

|  | Total TMB Model |  |  | UV TMB Model |  |  | Non-UV TMB Model |  |  | Joint Model |  |  |
|---|---|---|---|---|---|---|---|---|---|---|---|---|
|  | OR | 95% CI | p-value | OR | 95% CI | p-value | OR | 95% CI | p-value | OR | 95% CI | p-value |
| log Total TMB | 0.69 | 0.56, 0.84 | <0.001 | - | - | - | - | - | - | - | - | - |
| log UV TMB | - | - | - | 0.69 | 0.57, 0.84 | <0.001 | - | - | - | 0.70 | 0.54, 0.91 | 0.008 |
| log non-UV TMB | - | - | - | - | - | - | 0.73 | 0.57, 0.93 | 0.01 | 0.97 | 0.70, 1.35 | 0.900 |
| Covariates: |  |  |  |  |  |  |  |  |  |  |  |  |
| Age at Dx | 1.03 | 1.01, 1.04 | <0.001 | 1.03 | 1.01, 1.04 | <0.001 | 1.02 | 1.01, 1.04 | <0.001 | 1.03 | 1.01, 1.04 | <0.001 |
| Sex |  |  | 0.006 |  |  | 0.004 |  |  | 0.010 |  |  | 0.004 |
| Male | Ref. | - |  | Ref. | - |  | Ref. | - |  | Ref. | - |  |
| Female | 0.56 | 0.37, 0.85 |  | 0.55 | 0.36, 0.83 |  | 0.59 | 0.39, 0.88 |  | 0.55 | 0.36, 0.83 |  |
| Primary Site |  |  | <0.001 |  |  | <0.001 |  |  | <0.001 |  |  | <0.001 |
| Head/Neck | Ref. | - |  | Ref. | - |  | Ref. | - |  | Ref. | - |  |
| Trunk | 0.49 | 0.27, 0.88 |  | 0.48 | 0.27, 0.86 |  | 0.52 | 0.29, 0.92 |  | 0.48 | 0.27, 0.86 |  |
| Arms | 0.32 | 0.17, 0.59 |  | 0.32 | 0.17, 0.59 |  | 0.33 | 0.17, 0.60 |  | 0.32 | 0.17, 0.59 |  |
| Legs | 0.31 | 0.16, 0.57 |  | 0.30 | 0.16, 0.57 |  | 0.35 | 0.19, 0.65 |  | 0.30 | 0.16, 0.57 |  |
| Breslow Thickness |  |  | <0.001 |  |  | <0.001 |  |  | <0.001 |  |  | <0.001 |
| 1.1-2.0 mm | Ref. | - |  | Ref. | - |  | Ref. | - |  | Ref. | - |  |
| 2.1-4.0 mm | 1.87 | 0.99, 3.64 |  | 1.86 | 0.98, 3.61 |  | 1.83 | 0.97, 3.55 |  | 1.86 | 0.98, 3.62 |  |
| >4.0 mm | 3.06 | 1.63, 5.95 |  | 3.03 | 1.62, 5.88 |  | 3.00 | 1.60, 5.80 |  | 3.03 | 1.62, 5.89 |  |
| Ulceration |  |  | <0.001 |  |  | <0.001 |  |  | <0.001 |  |  | <0.001 |
| Absent | Ref. | - |  | Ref. | - |  | Ref. | - |  | Ref. | - |  |
| Present | 2.54 | 1.72, 3.80 |  | 2.54 | 1.72, 3.80 |  | 2.49 | 1.69, 3.72 |  | 2.54 | 1.72, 3.80 |  |
| Lymph Nodes |  |  | <0.001 |  |  | <0.001 |  |  | <0.001 |  |  | <0.001 |
| N0 | Ref. | - |  | Ref. | - |  | Ref. | - |  | Ref. | - |  |
| N1 | 3.22 | 2.07, 5.06 |  | 3.25 | 2.09, 5.11 |  | 3.20 | 2.06, 5.01 |  | 3.24 | 2.09, 5.10 |  |
| N2 | 8.69 | 4.46, 17.7 |  | 8.53 | 4.37, 17.4 |  | 8.81 | 4.56, 17.80 |  | 8.54 | 4.37, 17.4 |  |
| N3 | 12.00 | 5.50, 29.1 |  | 11.80 | 5.37, 28.5 |  | 12.90 | 5.94, 31.20 |  | 11.8 | 5.39, 28.6 |  |
| TIL Grade |  |  | 0.004 |  |  | 0.004 |  |  | 0.004 |  |  | 0.004 |
| Absent | Ref. | - |  | Ref. | - |  | Ref. | - |  | Ref. | - |  |
| Non-brisk | 0.58 | 0.30, 1.10 |  | 0.58 | 0.30, 1.10 |  | 0.56 | 0.29, 1.07 |  | 0.58 | 0.30, 1.10 |  |
| Brisk | 0.10 | 0.02, 0.41 |  | 0.10 | 0.02, 0.41 |  | 0.10 | 0.02, 0.41 |  | 0.10 | 0.02, 0.41 |  |
| Mitoses |  |  | 0.083 |  |  | 0.093 |  |  | 0.084 |  |  | 0.091 |
| Absent | Ref. | - |  | Ref. | - |  | Ref. | - |  | Ref. | - |  |
| Present | 2.36 | 0.90, 7.04 |  | 2.3 | 0.88, 6.85 |  | 2.34 | 0.90, 6.88 |  | 2.32 | 0.88, 6.93 |  |
| Solar Elastosis |  |  | 0.015 |  |  | 0.018 |  |  | 0.008 |  |  | 0.017 |
| Absent | Ref. | - |  | Ref. | - |  | Ref. | - |  | Ref. | - |  |
| Mild/Moderate | 1.21 | 0.75, 1.97 |  | 1.21 | 0.75, 1.97 |  | 1.13 | 0.70, 1.81 |  | 1.21 | 0.75, 1.97 |  |
| Severe | 0.45 | 0.23, 0.88 |  | 0.46 | 0.23, 0.89 |  | 0.40 | 0.21, 0.76 |  | 0.46 | 0.23, 0.89 |  |
| Cannot assess | 1.11 | 0.58, 2.14 |  | 1.12 | 0.58, 2.17 |  | 0.99 | 0.52, 1.90 |  | 1.12 | 0.58, 2.17 |  |

Abbreviations: OR = Odds Ratio, CI = Confidence Interval, Ref. = Reference Category, TIL = Tumor Infiltrating Lymphocytes

## DISCUSSION

In this study, we investigated tumor mutational burden (TMB) in the InterMEL study by decomposing overall TMB into two components: UV-induced mutations and non-UV mutations. We find that higher UV TMB in stage II/III tumors is associated with reduced 5-year mortality, whereas non-UV TMB showed no clear association with mortality. These findings suggest that UV TMB may be a better predictor of melanoma-specific mortality than overall TMB. Our results are consistent with those of previous investigations of UV TMB^20,22,23^; however prior studies were limited to advanced stage tumors or those in the TCGA database^29^ (which contains a disproportionate number of very large melanomas). In contrast, InterMEL focuses on stage II and III primary melanomas, which are more representative of those encountered in routine clinical practice. To our knowledge, this is the first study to demonstrate prognostic relevance for UV TMB measured at time of diagnosis in stage II and III primary melanomas.

Additionally, we find that UV TMB and non-UV TMB are differently associated with the clinicopathologic presentation of stage II/III primary melanomas. We observe higher mutation burden in men and older patients, both of which are commonly reported^21,23,30,31^. While male tumors are known to be more highly mutated, our analysis shows that this difference is due primarily to greater counts of UV-induced mutations, while there is no clear sex difference in non-UV mutations. This finding is consistent with prior evidence suggesting that men experience greater cumulative UV exposure^32^ and may accumulate UV damage at lower exposures than women^33,34^. Interestingly, we find that solar elastosis is associated with increased numbers of both UV-induced and non-UV mutations after controlling for age and sex. We have previously reported the correlation of solar elastosis with total TMB in the InterMEL cohort^35^. Because solar elastosis reflects cumulative damage caused by sun exposure, its association with increased UV-induced mutation burden is expected. It is however more puzzling that solar elastosis is also associated with higher counts of non-UV mutations, suggesting the possibility that chronic sun exposure may correlate with broader mutational processes not captured by UV signatures alone. Finally, we observe that only non-UV TMB was associated with mitotic activity, which is consistent with the idea that the non-UV component of TMB contains the clock-like mutations resulting from cell replication^36^. While other studies have found a negative association between clock-like mutations and survival^20,22^, we did not differentiate clock-like mutations from other non-UV mutations, which might explain why we found no association of non-UV TMB with mortality, rather than a negative association.

Although the effect of TMB on improved survival is theoretically expected to operate through increased immune activation, we do not see an association of UV mutations with histologic TIL grade. We may have observed this lack of association for a few reasons. For one, conventional TIL grading may be too crude. For example, most tumors in the InterMEL study were scored in the intermediate (non-brisk) TIL grade. The non-brisk category may contain tumors with a range of meaningful variation in immune activity that are not easily identified through the simple three grade TIL framework. Alternatively, UV-derived neoantigens could influence immune function in ways that are not directly proxied by quantities or locations of immune cells. It is possible that UV-induced mutations generate a more immunogenic repertoire of tumor antigens; however, neoantigen burden and immune recognition were not directly assessed in this study. Finally, it may be that the neoantigen hypothesis does not explain the mechanism of association between UV TMB and survival. Instead, UV TMB might be associated with a different favorable biological process that is un-related to immune infiltration.

There is great interest in identifying biomarkers that can differentiate patients at high risk of mortality from those with a better chance of responding to treatment. Prior studies suggest that TMB may predict response to treatment, although results have been mixed, leading to hypotheses that subsets of mutations (such as UV-induced mutations) may be more predictive^11,14,15,22,23,37,38^.

The focus on treatment response as a primary endpoint is biologically plausible since mutation counts are thought to proxy neoantigen load, potentially influencing tumor recognition by the immune system—and responsiveness to immune checkpoint therapy^6^. To better reflect the natural history of melanoma, our analysis was conducted in early-stage primary melanoma patients who did not receive modern adjuvant treatments. Thus, the endpoint for our analysis is not treatment response, but rather melanoma-specific mortality. Interestingly, while prior studies have linked UV TMB to treatment response, our findings suggest that higher UV TMB is associated with improved melanoma-specific survival even in the absence of contemporary adjuvant therapy. Future studies may be able to assess potential functional mechanisms, such as natural immune system involvement, underlying this association.

Our study has some limitations. Our UV and non-UV mutational signatures are inferred from panel-based sequencing, which means we are likely analyzing a subset of mutations in the tumor. Our results could differ if we had all observable mutations from whole-exome or whole-genome sequencing. The case-control design means that we are not observing population-based frequencies of mutations or outcomes. Further, this is an observational study, which precludes us from inferring causality in the relationship between mutations and disease outcomes. The InterMEL study is, to our knowledge, uniquely positioned to evaluate the prognostic significance of mutational burden in stage II/III melanoma because it combines centrally reviewed pathology, detailed clinical annotation, and high-quality molecular profiling. The dataset includes stage II and III primary melanomas, comprehensive clinicopathologic variables, and mutation measurements generated using a panel commonly employed in clinical practice. Together, these features provide a robust framework for evaluating the relative importance of UV and non-UV mutations, and here reveal that it is specifically the UV-induced component of TMB that associates with improved melanoma-specific survival.

## Data Availability

All data used in this analysis are available on dbGaP, at accession number phs003099.v2.p1

https://dbgap.ncbi.nlm.nih.gov/study/phs003099.v2.p1/#study

## Data Availability

All data used in this analysis are publicly available on dbGAP under accession number phs003099.v1.p1.

## Author Contributions

*Concept and Design*: Reynolds, Luo, Berwick. *Acquisition, Analysis, or Interpretation of Data*: All authors. *Drafting of the Manuscript*: Reynolds, Luo, Berwick. *Critical Review of the Manuscript for Important Intellectual Content*: All authors. *Statistical Analysis*: Reynolds, Luo. *Funding Acquisition*: Berwick, Thomas, Hernando, Shen, Begg, Gorlov, Amos, Orlow. *Administrative, Technical, or Material Support:* Boyce, Busam, Kenney, Sadeghi, Lee. This study was conducted by the InterMEL consortium, which includes the following study centers and personnel: Coordinating Center, University of New Mexico, Albuquerque, NM, USA: Marianne Berwick (Principal Investigator (MPI, contact PI), Li Luo (Investigator), Christopher I. Amos (Advisor), Tawny W. Boyce (Data Manager), Adam Z. Reynolds (Data Analyst), Dakai Zhu (Data Analyst), Charles Wiggins (Director, New Mexico Tumor Registry); New York University, Langone Cancer Center, New York, NY: Eva Hernando (PI-Project 3), Iman Osman (co-Investigator), Douglas Hanniford (Instructor), Diana Argibay (Laboratory Technician), Una Moran (Data Technician), for the Genomics Technology Center: Adriana Heguy (Director), Sitharam Ramaswami (Associate Research Scientist); Memorial Sloan Kettering Cancer Center, New York, NY: Ronglai Shen (PI-Project 4), Colin B. Begg (Biostatistician), Arshi Arora (Biostatistician), Venkatraman Seshan (Biostatistician), Allie Reiner (Assistant Research Biostatistician), Caroline E. Kostrzewa (Assistant Research Biostatistician), Klaus J. Busam (PI-Core 2), Irene Orlow (Co-PI-Core 2), Cecilia Lezcano (Dermatopathologist), Jessica M. Kenney (Research Assistant/Sr. Laboratory Specialist), Keimya D. Sadeghi (Research Assistant and Data Engineer), Kelli O’Connell (Research Biostatistician); initial optimization assays were performed at MSK by Heta Parmar, Siok Leong, and Sergio Corrales (Research Technicians); University of North Carolina, Chapel Hill, NC: Nancy E. Thomas (MPI), Kathleen Conway (Investigator), Sharon N. Edmiston (Research Analyst), David W. Ollila (Surgical Oncologist and Investigator), Honglin Hao (Laboratory Specialist), Eloise Parrish (Laboratory Specialist), Paul B. Googe (Dermatopathologist), Stergios J. Moschos (Oncologist), David Corcoran (Bioinformatician), Lan Lin (Data-base Manager); Melanoma Institute Australia, Sydney, Australia: Richard A. Scolyer (Dermato-pathologist and Site PI), Anne E. Cust (Epidemiologist and Site PI), James S. Wilmott (Scientist), Graham J. Mann (Cancer Geneticist and Former Site PI), Ping Shang (Scientist), Hazel Burke (Data Manager), Peter M. Ferguson (Pathologist), Valerie Jakrot (Research Manager); British Columbia Cancer Research Center, Vancouver, Canada: Tim K. Lee (Data Coordinator); Baylor College of Medicine, Houston, TX: Ivan P. Gorlov (PI-Core 3); Roswell Park Comprehensive Cancer Center, Buffalo, NY: Marc Ernstoff (Advisor), Paul N. Bogner (Dermatopathologist); The University of Texas MD Anderson Cancer Center, Houston, TX: Jeffrey E. Lee (Site PI), Melanoma Core (Technical Assistance); Dartmouth Cancer Center, Lebanon, NH: Judy R. Rees (Site PI), Shaofeng Yan (Dermato-pathologist); Case Western University, Cleveland, OH: Meg R. Gerstenblith (Site PI), Cheryl Thompson (Co-Investigator); Cleveland Clinic, Cleveland, OH: Jennifer S. Ko (Dermatopathologist and Site PI), Pauline Funchain (Co-Investigator), Peter Ngo (Dermatology Fellow); Yale University Cancer Center, New Haven, CT: Marcus Bosenberg (Site PI), Bonnie E. Gould Rothberg (Former Site PI), Gauri Panse (Dermatopathologist); Columbia University Medical School, New York, NY: Yvonne M. Saenger (Site PI), Benjamin T. Fullerton (Laboratory Technician); Huntsman Cancer Institute, Salt Lake City, UT: Sheri L. Holmen (Site PI), Howard Colman (Co-Investigator), Elise K. Brunsgaard (Clinical Fellow), David Wada (Dermatopathologist); Instituto Valenciano de Oncologia, Valencia, Spain: Eduardo Nagore (Site PI), Esperanza Manrique-Silva (Dermatologist), Celia Requena (Dermatologist), Victor Traves (Pathologist), David Millan-Esteban (Post-Doctoral Fellow); Patient Advocate: Michelle Rainka.

